# Longitudinal Physiological Data from a Wearable Device Identifies SARS-CoV-2 Infection and Symptoms and Predicts COVID-19 Diagnosis

**DOI:** 10.1101/2020.11.06.20226803

**Authors:** Robert P. Hirten, Matteo Danieletto, Lewis Tomalin, Katie Hyewon Choi, Micol Zweig, Eddye Golden, Sparshdeep Kaur, Drew Helmus, Anthony Biello, Renata Pyzik, Ismail Nabeel, Alexander Charney, Benjamin Glicksberg, Matthew Levin, David Reich, Dennis Charney, Erwin P Bottinger, Laurie Keefer, Mayte Suarez-Farinas, Girish N. Nadkarni, Zahi A. Fayad

**Affiliations:** The Dr. Henry D. Janowitz Division of Gastroenterology, Icahn School of Medicine at Mount Sinai, New York, NY, USA; The Hasso Plattner Institute for Digital Health at the Mount Sinai, New York, NY, USA; Department of Genetics and Genomic Sciences, Icahn School of Medicine at Mount Sinai, New York, NY, USA; Center for Biostatistics, Department of Population Health Science and Policy, Icahn School of Medicine at Mount Sinai; The BioMedical Engineering and Imaging Institute, Icahn School of Medicine at Mount Sinai, New York, NY, USA; The Department of Psychiatry, Icahn School of Medicine at Mount Sinai, New York, NY, USA; The Pamela Sklar Division of Psychiatric Genomics, Icahn School of Medicine at Mount Sinai, New York, NY, USA; Department of Anesthesiology, Perioperative and Pain Medicine, Icahn School of Medicine at Mount Sinai; Department of Environmental Medicine and Public Health, Icahn School of Medicine at Mount Sinai, New York, NY, USA; Office of the Dean, Icahn School of Medicine at Mount Sinai; The Department of Medicine, Icahn School of Medicine at Mount Sinai, New York, NY, USA; The Charles Bronfman Institute for Personalized Medicine, Icahn School of Medicine at Mount Sinai, New York, NY, USA; Department of Diagnostic, Molecular and Interventional Radiology, Icahn School of Medicine at Mount Sinai; Nash Family Department of Neuroscience, Icahn School of Medicine at Mount Sinai

**Author notes:** **Correspondence:** Robert P Hirten MD, 1468 Madison Avenue, Annenberg Building RM 5-12, New York, NY 10029.

## Abstract

**Background:** Changes in autonomic nervous system function, characterized by heart rate variability (HRV), have been associated with and observed prior to the clinical identification of infection. We performed an evaluation of this metric collected by wearable devices, to identify and predict Coronavirus disease 2019 (COVID-19) and its related symptoms.

**Methods:** Health care workers in the Mount Sinai Health System were prospectively followed in an ongoing observational study using the custom Warrior Watch Study App which was downloaded to their smartphones. Participants wore an Apple Watch for the duration of the study measuring HRV throughout the follow up period. Survey’s assessing infection and symptom related questions were obtained daily.

**Findings:** Using a mixed-effect COSINOR model the mean amplitude of the circadian pattern of the standard deviation of the interbeat interval of normal sinus beats (SDNN), a HRV metric, differed between subjects with and without COVID-19 (p=0.006). The mean amplitude of this circadian pattern differed between individuals during the 7 days before and the 7 days after a COVID-19 diagnosis compared to this metric during uninfected time periods (p=0.01). Significant changes in the mean MESOR and amplitude of the circadian pattern of the SDNN was observed between the first day of reporting a COVID-19 related symptom compared to all other symptom free days (p=0.01).

**Interpretation:** Longitudinally collected HRV metrics from a commonly worn commercial wearable device (Apple Watch) can identify the diagnosis of COVID-19 and COVID-19 related symptoms. Prior to the diagnosis of COVID-19 by nasal PCR, significant changes in HRV were observed demonstrating its predictive ability to identify COVID-19 infection.

**Funding:** Support was provided by the Ehrenkranz Lab For Human Resilience, the BioMedical Engineering and Imaging Institute, The Hasso Plattner Institute for Digital Health at Mount Sinai, The Mount Sinai Clinical Intelligence Center and The Dr. Henry D. Janowitz Division of Gastroenterology.

## INTRODUCTION

Coronavirus disease 2019 (COVID-19) has resulted in over 41 million infections and more than 1.1 million deaths.^1^ A prolonged incubation period and variable symptomatology has facilitated disease spread, with approximately 30-45% of individuals having asymptomatic SARS-CoV-2 infections, and testing generally limited to only symptomatic individuals.^2-4^ Health care workers (HCWs), characterized as any type of worker in a health care system, represent a vulnerable population with a threefold increased risk of infection compared to the general population.^5^ This increased risk of transmission is important in healthcare settings, where asymptomatic or pre-symptomatic HCWs can shed the virus contributing to transmission within healthcare facilities and their households.^6^

Digital health technology offers an opportunity to address the limitations of traditional public health strategies aimed at curbing COVID-19 spread.^7^ Smart phone Apps are effective in using symptoms to identify those possibly infected with SARS-CoV-2, but they rely on ongoing participant compliance and self-reported symptoms.^8^ Wearable devices are commonly used for remote sensing and provide a means to objectively quantify physiological parameters including heart rate, sleep, activity and measures of autonomic nervous system (ANS) function (e.g., heart rate variability [HRV]).^9^ The addition of physiological data from wearable devices to symptom tracking Apps has been shown to increase the ability to identify those infected with SARS-CoV-2.^10^

HRV is a physiological metric providing insight into the interplay between the parasympathetic and sympathetic nervous system which modulate cardiac contractility and cause variability in the beat-to-beat intervals.^11^ It exhibits a 24 hour circadian pattern with relative sympathetic tone during the day and parasympathetic activity at night.^12-14^ Changes in this circadian pattern can be leveraged to identify different physiological states. Several studies have demonstrated that lower HRV, indicating increased sympathetic balance, is a reliable predictor of infection onset.^15,16^ However, HRV and its dynamic changes over time have not been evaluated as a marker or predictor of COVID-19. In response to the COVID-19 pandemic we launched The Warrior Watch Study™, employing a novel smartphone App to remotely enroll and monitor HCWs throughout the Mount Sinai Health System in New York City, a site of initial case surge. This digital platform enables remote survey delivery to Apple iPhones and passive collection of Apple Watch data, including HRV. The aim of this study is to determine if SARS-CoV-2 infections can be identified and predicted prior to a positive test result using the longitudinal changes in HRV metrics derived from the Apple Watch.

## METHODS

### Study Design

The primary aim of the study was to determine whether changes in HRV can differentiate participants infected or not infected with SARS-CoV-2. The secondary aim was to see if changes in HRV can predict the development of a SARS-CoV-2 infection prior to diagnosis by a SARS-CoV-2 nasal PCR. Exploratory aims were (1) to determine whether changes in HRV can identify the presence of COVID-19 related symptoms; (2) to determine whether changes in HRV can predict the development of COVID-19 related symptoms; and (3) to evaluate how HRV changed throughout the infection and symptom period.

HCWs in the Mount Sinai Health System were enrolled in an ongoing prospective observational cohort study. Eligible participants were ≥18 years of age, current employees in the Mount Sinai Health System, had an iPhone Series 6 or higher, and had or were willing to wear an Apple Watch Series 4 or higher. Participants were excluded if they had an underlying autoimmune disease or were on medications known to interfere with ANS function. A positive COVID-19 diagnosis was defined as a positive SARS-CoV-2 nasal PCR swab reported by the participant. Daily symptoms were collected including fevers/chills, tired/weak, body aches, dry cough, sneezing, runny nose, diarrhea, sore throat, headache, shortness of breath, loss of smell or taste, itchy eyes, none, or other. This study was approved by the Institutional Review Board at The Icahn School of Medicine at Mount Sinai.

### Study Procedures

Participants downloaded the custom Warrior Watch App to complete eligibility questionnaires and sign an electronic consent form. Participants completed an App-based baseline assessment collecting demographic information, prior COVID-19 diagnosis history, occupation, and medical history and were then followed prospectively through the App. Daily survey questionnaires captured COVID-19 related symptoms, symptom severity, SARS-CoV-2 nasal PCR results, serum SARS-CoV-2 antibody test results, and daily patient care related exposure (**Supplementary Table 1**). Participants carried out their normal activities throughout the study and were instructed to wear the Apple Watch for a minimum duration of 8 hours per day.

### Wearable Monitoring Device and Autonomic Nervous System Assessment

HRV was measured via the Apple Watch Series 4 or 5, which are commercially available wearable devices. Participants wore the device on the wrist and connected it via Bluetooth to their iPhone. The Watch is equipped with an enhanced photoplethysmogram (PPG) optical heart sensor that combines a green LED light paired with a light sensitive photodiode generating time series peaks that correlate with the magnitude of change in the green light generated from each heartbeat.^17^ Data are filtered for ectopic beats and artifact. The time difference between heartbeats is classified as the Interbeat Interval (IBI) from which HRV is calculated. The Apple Watch and the Apple Health app automatically calculate HRV using the standard deviation of the IBI of normal sinus beats (SDNN), measured in milliseconds (ms). This time domain index reflects both sympathetic and parasympathetic nervous system activity and is calculated by the Apple Watch during ultra-short-term recording periods of approximately 60 seconds.^11^ The Apple Watch generates several HRV measurements throughout a 24-hour period. HRV metrics are stored in a locally encrypted database accessible through the iPhone Health app which is retrieved through our custom Warrior Watch App. Data is transferred from the iPhone and Apple Watch upon completion of the e-consent and any survey in the App. Wearable data is stored locally allowing retrieval during the days when surveys are not completed by participants.

### Statistical Analysis

#### Heart Rate Variability Modelling

The HRV data collected through the Apple Watch was characterized by a circadian pattern, a sparse sampling over a 24-hour period, and a non-uniform timing across days and participants. These characteristics bias easily derived features including mean, maximum and minimum creating the need to derive methods that model the circadian rhythm of HRV. A COSINOR model was used to model daily circadian rhythm over a 24 hour period with the non-linear function Y(t) = M+*Acos*(2*π*t/*τ* + *ϕ*) + e_i_(t) [equation 1], where τ is the period (*τ* =24h), M is the Midline Statistic of Rhythm (MESOR), a rhythm-adjusted mean, A is the amplitude, a measure of half the extent of variation within a day and F is the Acrophase, a measure of the time of overall high values recurring in each day (**Supplementary Figure 1**). This non-linear model with 3 parameters has the advantage of being easily transformed into a linear model by recoding time (t) into two new variables x and z as *x* = sin(2*π*t/*τ*), *z* = sin(2*π*t/*τ*). HRV can then be written as Y(t)=M+*β*x_t_ + *γ*z_t_ + e_i_(t) [equation 2], where the linear coefficients *β, γ* of the linear model in equation 2 are related to the non-linear parameters of the non-linear model in equation 1 by *β* = *Acos*(*ϕ*) *γ* = −*Asin*(*ϕ*). One can estimate the linear parameters *β, γ* and then obtain the A and *ϕ* as:

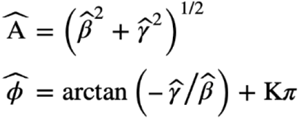

We took advantage of the longitudinal structure of the data to identify a participant specific daily pattern and then measured departures from this pattern as a function of COVID-19 diagnosis or other relevant covariates. In order to do so we used a mixed-effect COSINOR model, where the HRV measure of participant i at time t can be written as HRV_it_ = (M+*β*.x_it_ + *γ*.z_it_) + *W*_*it*_. *θ*_*i*_+ e_i_(t), e_i_(t)∼N(0,s), and where M, *β* and *γ* are the population parameters (fixed-effects) and *θ*_i_ is a vector of random effects and assumed to follow a multivariate normal distribution *θ* _i_∼N(0,Σ). In this context the introduction of random effects intrinsically model the correlation due to the longitudinal sampling. To measure the impact of any covariate C on the participants’ daily curve, we can introduce such covariates as fixed-effects as its interactions with x and z: HRV_it_ = M+*a*_o_C_i_+(*β* + *a*_2_C_i_).x_it_ + (*γ* + *a* _3_C_i_.)z_it_ + *W*_*it*_. *θ*_*i*_ + e_i_(t) [equation 3]. Model parameters and the standard errors of equation 3 can be estimated via maximum likelihood or reweighted least squares (REWL) and hypothesis testing can be carried out for any comparison that can be written as a linear function of *a*′*s, β and γ* parameters.

However, to test if the COSINOR curve, defined by the non-linear parameters M, A and *ϕ* in equation 1 differs between the populations defined by the covariate C, we proposed the following bootstrapping procedure where for each resampling iteration we: (1) Fit a linear mixed-effect model using REWL; (2) Estimated the marginal means obtaining the linear parameters for each group defined by covariate C; (3) Used the inverse relationship to estimate marginal means M, A and *ϕ* for each group defined by C; and (4) Defined the bootstrapping statistics as the pairwise differences of M, A and *ϕ* between groups defined by C. For such iterations, the confidence intervals for the non-linear parameter was defined using standard bootstrap techniques, as well deriving the p-values for the differences of each non-linear parameter between groups defined by Ci. Age and sex were included as a covariate in HRV analyses and admitted invariant and time-variant covariates.

#### Association and Prediction of COVID-19 Diagnosis and Symptoms

The relationship between a COVID-19 diagnosis and change in HRV curves were evaluated. To test this association, we defined the time variant covariate C_it_ for participant i at time t as:

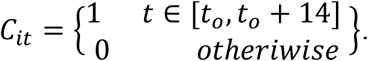

HRV metrics for the 14 days following the time of first positive SARS-CoV-2 nasal PCR test were used to define the positive SARS-CoV-2 infection window. To evaluate the predictive ability of changes in HRV prior to a COVID-19 diagnosis and to explore its changes during the infection period, the time variant covariate was used to characterize the following 4 groups: healthy uninfected individuals [t<t_0_-7], 7 days before COVID-19 diagnosis [t≥t_0_-7, t<t_0_], the first 7 days post COVID-19 diagnosis [t_0_≤t<t_0_+7] and the 7-14 days post diagnosis [t_0_+7≤t<t_0_+14].

To determine the association between COVID-19 symptoms and changes in HRV metrics, we defined being symptomatic as the 1^st^ day of a reported symptom and compared this to all other days. To evaluate the predictive ability of HRV to identify upcoming symptom days and to explore its changes over time the time variant covariate was used to characterize the following 4 groups: healthy asymptomatic individuals for t<t_0_-1, one day before COVID-19 symptoms [t≥t_0_-1, t<t_0_], the first day of COVID-19 symptoms [t_0_≤t<t_0_+1] and one day post COVID-19 symptom development [t_0_+1≤t<t_0_+2].

## RESULTS

Two hundred and ninety-seven participants were enrolled between April 29^th^ and September 29^th^, 2020, when data was censored for analysis (**Table 1**). The median age at enrollment was 36 years with 69% of participants being women. Twenty participants reported having a positive SARS-CoV-2 nasal PCR test prior to enrollment, while 28 participants reported having a positive blood antibody test prior to joining the study. The median duration of follow up was 42 days (range 0-152 days). A median of 28 HRV samples (range 1-129) were obtained per participant. Study compliance over the follow up period, defined as participants answering over 50% of daily surveys, was 70.4%.

**Table 1.** Baseline demographics of participants at enrollment.

|  | **Cohort (n=297)** |
| --- | --- |
| Age, mean (SD) | 36.3 (9.8) |
| Body Mass Index, mean (SD) | 25.6 (5.7) |
| Female Gender (%) | 204 (69.4) |
| Race (%) |  |
| Asian | 73 (24.6) |
| Black | 29 (9.8) |
| Other | 43 (14.5) |
| White | 108 (36.4) |
| Ethnicity (%) |  |
| Hispanic | 44 (14.8) |
| Baseline Positive SARS-CoV-2 nasal PCR (%) | 20 (6.7) |
| Baseline Positive SARS-CoV-2 serum antibody (%) | 28 (9.4) |
| Occupation* (%) |  |
| Clinical non-Trainee | 198 (68.0) |
| Clinical Trainee | 36 (12.4) |
| Non-clinical Staff | 57 (19.6) |
| Baseline Smoking Status (%) |  |
| Current/Past smoker | 35 (11.9) |
| Never/Rarely smoker | 259 (88.1) |
| Baseline Immune Suppressing Medication (%) | 4 (1.4) |

PCR, polymerase chain reaction; SD, standard deviation
\*Clinical trainee defined as a resident or fellow; clinical non-trainee defined as HCWs reporting at least one patient facing day during follow up, exclusive of resident and fellows; non-clinical staff defined as a HCW who did not report a patient facing day during follow up.

### Identification and Prediction of COVID-19 Diagnosis

Thirteen participants reported a positive SARS-CoV-2 nasal PCR during the follow up period. The mean MESOR, acrophase and amplitude of the circadian SDNN pattern in participants diagnosed with and without COVID-19 are described in **Table 2**. A significant difference in the circadian pattern of SDNN was observed in participants diagnosed with COVID-19 compared to those without COVID-19. There was a significant difference (p=0.006) between the mean amplitude of SDNNs circadian pattern in those with (1.23 ms, 95% CI −1.94-3.11) and without COVID-19 (5.30 ms, 95% CI 4.97-5.65). No difference was observed between the MESOR (p=0.46) or acrophase (p=0.80) in these two infection states (**Figure 1a-c**).

**Table 2.** HRV parameters in participants with and without COVID-19 diagnosed based on SARS-CoV-2 nasal PCR swabs.

| Parameter | Parameter Mean, ms (95% CI)<br>COVID-19 Negative | Parameter Mean, ms (95% CI)<br>COVID-19 Positive | Difference<br>(95% CI) | p-value |
| --- | --- | --- | --- | --- |
| MESOR | 43.57 (41.40-45.40) | 42.46 (38.90-45.79) | -1.12 (-4.22- 1.73) | 0.46 |
| Amplitude | 5.30 (4.97-5.65) | 1.23 (-1.94- 3.11) | -4.07 (-7.29- -2.07) | **0.006** |
| Acrophase | -2.44 (-2.49- -2.39) | -2.23 (-2.22- -4.24) | 0.22 (-1.74- 2.43) | 0.80 |

**Figure 1:**
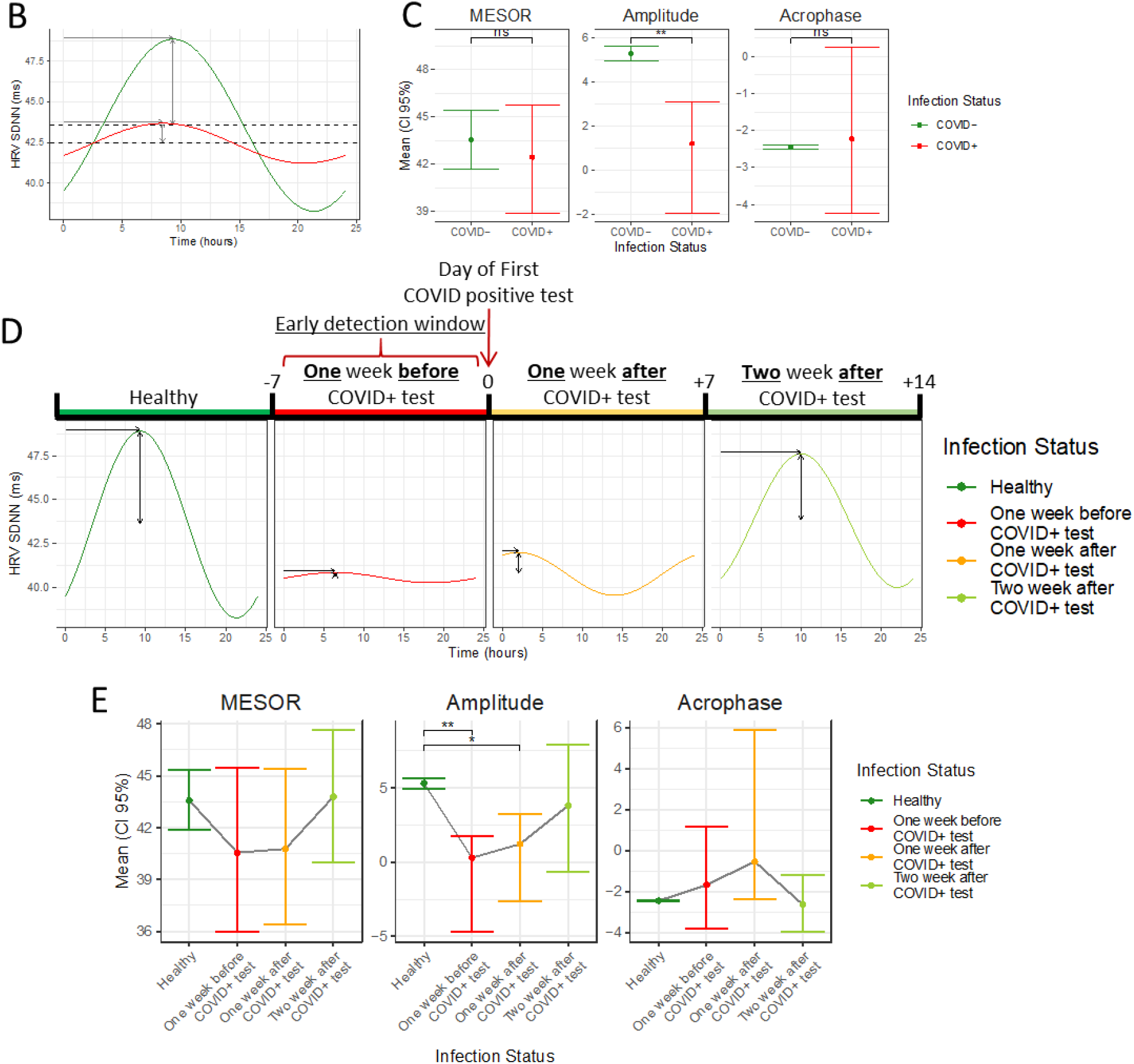
Relationship between HRV circadian rhythm and COVID-19 status. Timeline (A) illustrates HRV measures from the time of COVID-19 diagnosis via nasal PCR and during the following 2 weeks where subjects were deemed to be COVID-19+ (red), and were compared with measurements outside this window, where subjects were deemed COVID- (green). Daily HRV rhythm (B) during days with COVID+ (red) and COVID- (green) diagnosis, time (hours) is indicated by the x-axis while SDNN (ms) is indicated by the y-axis. Plots (C) showing Mean and 95%CI for the parameters defining the circadian rhythm: Acrophase, Amplitude and MESOR in COVID+ (red) and COVID- (green) days. Daily HRV pattern (D) for days were subjects were healthy (green), 7 days before COVID-19+ test (red), 7 days after COVID-19+ test (orange) and 7-14 days after COVID-19+ test (light green), time (hours) is indicated by the x-axis while HRV (ms) is indicated by the y-axis. Mean and 95% CI for the Acrophase, Amplitude and MESOR of the HRV measured on days were participants were Healthy (green), 7 days before COVID-19+ test (red), 7 days after COVID-19+ test (orange) and 7-14 days after COVID-19+ test (light green). **+**p<0.1;*p<0.05; **p<0.01; ***p<0.001; ns, not significant

The mean MESOR, acrophase and amplitude of the circadian SDNN pattern for those without COVID-19, those during the 7 days prior to a COVID-19 diagnosis, participants during the 7 days after a COVID-19 diagnosis and those during the 7-14 days after a COVID-19 diagnosis are described in **Table 3**. Significant changes in the circadian pattern of SDNN were observed in participants during the 7 days prior and the 7 days after a diagnosis of COVID-19 when compared to uninfected participants. There was a significant difference between the amplitude of the SDNN circadian rhythm between uninfected participants (5.31 ms, 95% CI 4.95-5.67) compared to individuals during the 7 day period prior to a COVID-19 diagnosis (0.29 ms, 95% CI −4.68-1.73; p=0.01) and participants during the 7 days after a COVID-19 diagnosis (1.22 ms, 95% CI −2.60-3.25; p=0.01). There were no other significant differences between the MESOR, amplitude, and acrophase of SDNNs circadian rhythm observed between healthy individuals, individuals 7 days before a COVID-19 diagnosis, individuals 7 days after a COVID-19 diagnosis, and individuals 7-14 days after infection (**Figure 1d-e**).

**Table 3.** Comparison of HRV parameters based on the time period before and after diagnosis.

| Parameter | Period Around<br>COVID-19<br>Diagnosis | Mean, ms<br>(95% CI) | Period Around<br>COVID-19<br>Diagnosis | Mean, ms<br>(95% CI) | Difference<br>(95% CI) | p-value |
| --- | --- | --- | --- | --- | --- | --- |
| MESOR | 7 Days Before | 40.56<br>(35.98-45.46) | Uninfected | 43.58<br>(41.88-45.37) | -3.03<br>(-6.98-1.02) | 0.13 |
|  | 7 Days After | 40.77<br>(36.44-45.42) | Uninfected | 43.58<br>(41.88-45.37) | -2.81<br>(-6.73-1.10) | 0.17 |
|  | 7-14 Days After | 43.80<br>(40.01-47.65) | Uninfected | 43.58<br>(41.88-45.37) | 0.22<br>(-3.39- 3.73) | 0.89 |
|  | 7 Days Before | 40.56<br>(35.98-45.46) | 7-14 Days After | 43.80<br>(40.01-47.65) | -3.24<br>(-9.63-3.33) | 0.32 |
|  | 7 Days After | 40.77<br>(36.44-45.42) | 7-14 Days After | 43.80<br>(40.01-47.65) | -3.03<br>(-6.98-1.02) | 0.13 |
|  | 7 Days After | 40.77<br>(36.44-45.42) | 7 Days Before | 40.56<br>(35.98-45.46) | 0.217<br>(-3.39-3.73) | 0.89 |
| Amplitude | 7 Days Before | 0.29<br>(-4.68-1.73) | Uninfected | 5.31<br>(4.95-5.67) | -5.02<br>(-10.14- -3.58) | **0.01** |
|  | 7 Days After | 1.22<br>(-2.60-3.25) | Uninfected | 5.31<br>(4.95-5.67) | -4.09<br>(-7.87- -1.93) | **0.01** |
|  | 7-14 Days After | 3.80<br>(-0.64-7.88) | Uninfected | 5.31<br>(4.95-5.67) | -1.51<br>(-5.79-2.35) | 0.48 |
|  | 7 Days Before | 0.29<br>(-4.68-1.73) | 7-14 Days After | 3.80<br>(-0.64-7.88) | -3.51<br>(-10.50-0.22) | 0.20 |
|  | 7 Days After | 1.22<br>(-2.60-3.25) | 7-14 Days After | 3.80<br>(-0.64-7.88) | -2.58<br>(-8.44-2.08) | 0.34 |
|  | 7 Days After | 1.22<br>(-2.60-3.25) | 7 Days Before | 0.29<br>(-4.68-1.73) | 0.93<br>(-1.92- 5.83) | 0.58 |
| Acrophase | 7 Days Before | -1.67<br>(-3.78-1.19) | Uninfected | -2.44<br>(-2.49- -2.39) | 0.78<br>(-1.4- 3.62) | 0.45 |
|  | 7 Days After | -0.53<br>(-2.39-5.89) | Uninfected | -2.44<br>(-2.49- -2.39) | 1.92<br>(0.03-8.13) | 0.48 |
|  | 7-14 Days After | -2.63<br>(-3.95-1.19) | Uninfected | -2.44<br>(-2.49- -2.39) | -0.19<br>(-1.39-1.16) | 0.70 |
|  | 7 Days Before | -1.67<br>(-3.78-1.19) | 7-14 Days After | -2.63<br>(-3.95-1.19) | 0.96<br>(-1.85-4.32) | 0.55 |
|  | 7 Days After | -0.53<br>(-2.39-5.89) | 7-14 Days After | -2.63<br>(-3.95-1.19) | 2.10<br>(0.10-8.29) | 0.35 |
|  | 7 Days After | -0.53<br>(-2.39-5.89) | 7 Days Before | -1.67<br>(-3.78-1.19) | 1.14<br>(-1.34- 7.27) | 0.58 |

### Identification and Prediction of COVID-19 Symptoms

Symptoms were frequently reported during the follow up period with the greatest number of participants reporting feeling tired or weak (n=87), followed by headaches (n= 82) and sore throat (n=60) (**Table 4**). Evaluating the days when participants experienced symptoms, we found that loss of smell or taste were reported the most with a mean of 138 days. This was followed by feeling tired or weak, reported a mean of 25 days and runny nose, reported a mean of 19.5 days (**Figure 2**). The mean MESOR, acrophase and amplitude observed in the circadian SDNN pattern in participants on the first day a symptom and on all other days of follow up are reported in **Table 5**. There was a significant difference in the circadian SDNN pattern between participants on the first day a symptom was reported compared to all other days of follow up. Specifically, there was a significant difference (p=0.01) between the mean MESOR of SDNNs circadian pattern on the first day of symptoms (46.01 ms, 95% CI 43.37-48.77) compared to all other days (43.48 ms, 95% CI 41.77-45.27). Similarly, there was a significant difference (p=0.01) between the mean amplitude of SDNNs circadian pattern on the first day of symptoms (2.58 ms, 95% CI 0.26-5.00) compared to all other days (5.30 ms, 95% CI 4.95-5.66) (**Figure 3a-c**).

**Table 4.**
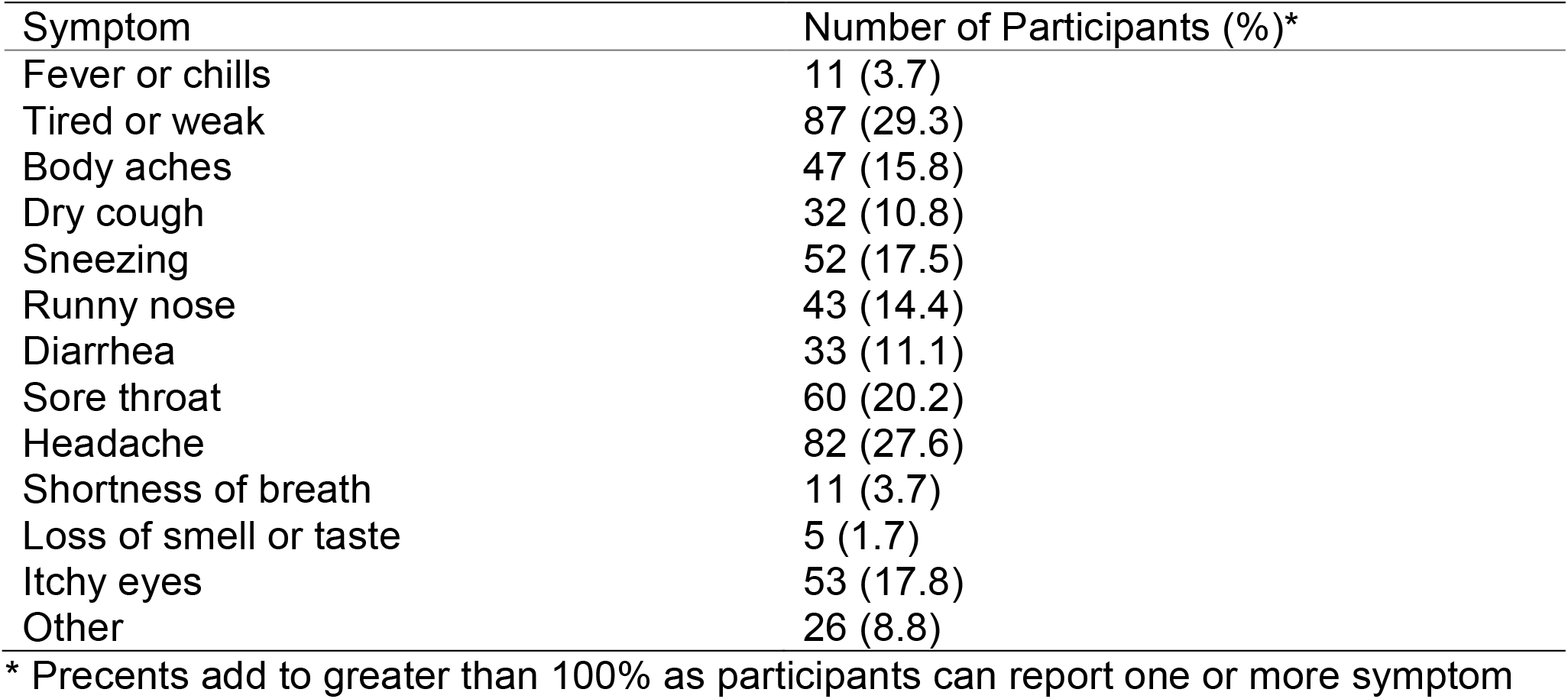
Number of participants reporting each symptom.

| Symptom | Number of Participants (%)<sup>*</sup> |
| --- | --- |
| Fever or chills | 11 (3.7) |
| Tired or weak | 87 (29.3) |
| Body aches | 47 (15.8) |
| Dry cough | 32 (10.8) |
| Sneezing | 52 (17.5) |
| Runny nose | 43 (14.4) |
| Diarrhea | 33 (11.1) |
| Sore throat | 60 (20.2) |
| Headache | 82 (27.6) |
| Shortness of breath | 11 (3.7) |
| Loss of smell or taste | 5 (1.7) |
| Itchy eyes | 53 (17.8) |
| Other | 26 (8.8) |

<sup>\*</sup> Percents add to greater than 100% as participants can report one or more symptom

**Table 5.** HRV parameters on the first day of reported symptoms compared to all other symptom free days.

| Parameter | Parameter Mean, ms (95% CI) First Day of Symptoms | Parameter Mean, ms (95% CI) All Other Days | Difference (95% CI) | p-value |
| --- | --- | --- | --- | --- |
| MESOR | 46.01 (43.37-48.77) | 43.48 (41.77-45.27) | 2.53 (0.82-4.36) | **0.01** |
| Amplitude | 2.58 (0.26-5.00) | 5.30 (4.95-5.66) | -2.73 (-5.16- 0.31) | **0.01** |
| Acrophase | -2.21 (-2.83- -1.58) | -2.44 (-2.49- -2.39) | 0.24 (-0.38- 0.88) | 0.44 |

**Figure 2.**
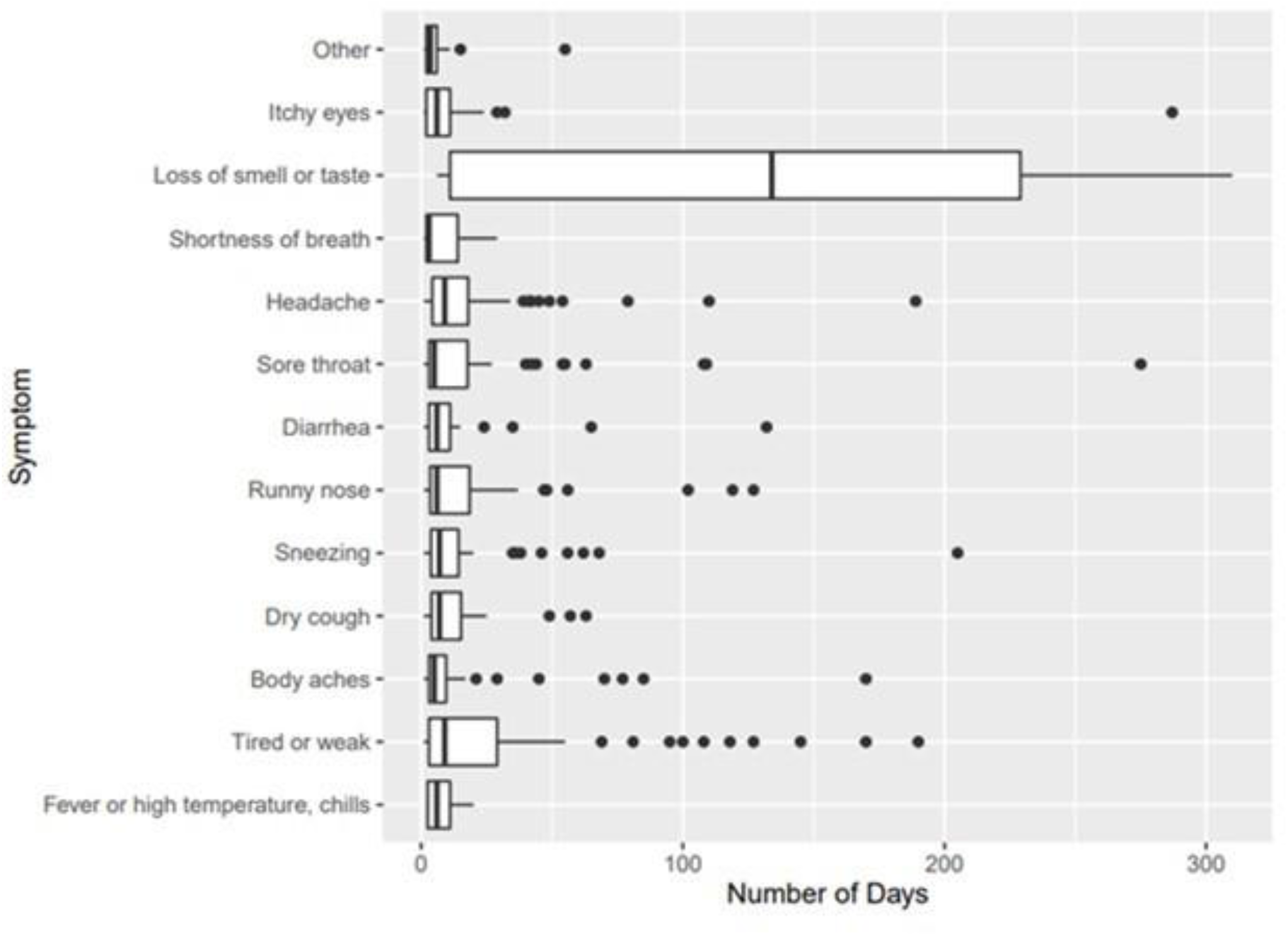
Number of symptom days per participant when evaluating days when participants reported symptoms.

**Figure 3:**
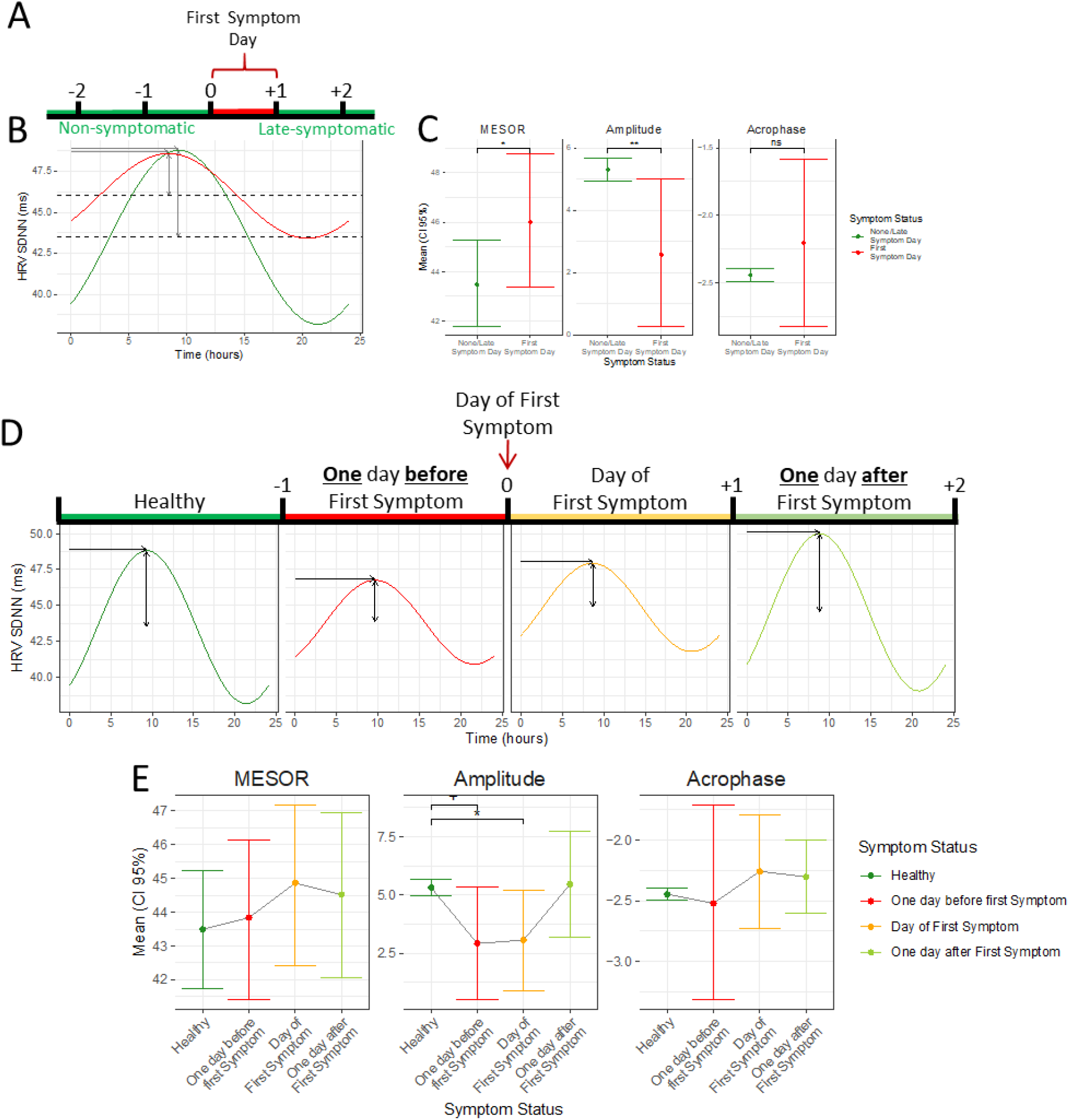
Relationship between HRV circadian rhythm and symptom onset. Timeline (A) illustrates timing of symptom onset, HRV profiles of the first-symptom day (red) were compared to all other days (green). Daily HRV rhythm (B) on day of first symptom (red) and non/late-symptom (green) days, time (hours) is indicated by the x-axis and HRV (ms) is indicated by the y-axis. Plots (C) showing Mean and 95%CI for the parameters defining the circadian rhythm: Acrophase, Amplitude and MESOR on first symptom (red) and non/late-symptom (green) days. Daily HRV pattern (D) for non/late-symptomatic days (green), the day before first symptom (red), day of first symptom (orange) and day after first symptom (light green), time (hours) is indicated by the x-axis while HRV (ms) is indicated by the y-axis. Mean and 95% CI for the Acrophase, Amplitude and MESOR of the HRV measured on non/late-symptomatic days (green), the day before first symptom (red), day of first symptom (orange) and day after first symptom (light green), **+**p<0.1;*p<0.05; **p<0.01; ***p<0.001; ns, not significant

The mean MESOR, acrophase and amplitude observed in the circadian SDNN pattern in participants on the day before symptoms develop, on the first day of the symptom, on the day following the first day of the symptom and on all other days are reported in **Table 6**. Significant changes in the circadian pattern of SDNN were observed, specifically in the mean amplitude (p=0.04) when comparing participants on the first day of the symptom (3.07 ms, 95% CI 0.88-5.22) to all other days (5.32 ms, 95% CI 4.99-5.66). Excluded from this analysis was the day prior and day after the first symptomatic day. Changes in SDNN characteristics trended toward significance prior to the development of symptoms. Specifically, the differences in the mean amplitude of SDNNs circadian pattern trended toward significance when comparing the day prior to symptom development (2.92 ms, 95% CI 0.50-5.33) with all other days (5.32, 95% CI 4.99-5.66; p=0.056). Again, excluded from the analysis was the first day of the symptom and the day after the first symptomatic day. Additionally, there was trend toward significance when comparing the amplitude of the SDNN circadian pattern between participants during the first day of the symptom (3.07 ms, 95% CI 0.88-5.22) with the one day after the first symptom was reported (5.47 ms, 95% CI 3.16-7.76; p=0.56). Excluded from the analysis was the day prior to symptom development and all other days. There were no other significant differences between the MESOR, amplitude, and acrophase of SDNNs circadian rhythm when comparing participants on the day before symptoms develop, on the first day of the symptom, on the day following the first day of the symptom and on all other days (**Figure 3d-e**).

**Table 6.** Comparison of HRV parameters based on symptom state and the time-period before and after the first day of reported symptoms.

| Parameter | Symptom State | Mean, ms<br>(95% CI) | Symptom State | Mean, ms<br>(95% CI) | Difference<br>(95% CI) | p-value |
| --- | --- | --- | --- | --- | --- | --- |
| MESOR | One day after 1<sup>st</sup> symptom day | 44.52<br>(42.05-46.94) | Asymptomatic | 43.49<br>(41.74-45.21) | 1.03<br>(-0.64-2.67) | 0.21 |
|  | One day before 1<sup>st</sup> symptom day | 43.84<br>(41.41-46.15) | Asymptomatic | 43.49<br>(41.74-45.21) | 0.34<br>(-1.46-2.23) | 0.73 |
|  | 1<sup>st</sup> day of symptom | 44.87<br>(42.42-47.18) | Asymptomatic | 45.49<br>(41.74-45.21) | 1.37<br>(-0.24-3.04) | 0.11 |
|  | One day before 1<sup>st</sup> symptom day | 43.84<br>(41.41-46.15) | One day after 1<sup>st</sup> symptom day | 44.52<br>(42.05-46.94) | -0.69<br>(-3.72-2.47) | 0.66 |
|  | 1<sup>st</sup> day of symptom | 44.87<br>(42.42-47.18) | One day after 1<sup>st</sup> symptom day | 44.52<br>(42.05-46.94) | 0.34<br>(-1.46-2.23) | 0.73 |
|  | 1<sup>st</sup> day of symptom | 44.87<br>(42.42-47.18) | One day before 1<sup>st</sup> symptom day | 43.84<br>(41.41-46.15) | 1.03<br>(-0.64-2.67) | 0.21 |
| Amplitude | One day after 1<sup>st</sup> symptom day | 5.47<br>(3.16-7.76) | Asymptomatic | 5.32<br>(4.99-5.66) | 0.15<br>(-2.21-2.37) | 0.91 |
|  | One day before 1<sup>st</sup> symptom day | 2.92<br>(0.50-5.33) | Asymptomatic | 5.32<br>(4.99-5.66) | -2.40<br>(-4.75- -0.07) | 0.056 |
|  | 1<sup>st</sup> day of symptom | 3.07<br>(0.88-5.22) | Asymptomatic | 5.32<br>(4.99-5.66) | -2.25<br>(-4.38- -0.27) | **0.04** |
|  | One day before 1<sup>st</sup> symptom day | 2.92<br>(0.50-5.33) | One day after 1<sup>st</sup> symptom day | 5.47<br>(3.16-7.76) | -2.55<br>(-6.64- 1.65) | 0.25 |
|  | 1<sup>st</sup> day of symptom | 3.07<br>(0.88-5.22) | One day after 1<sup>st</sup> symptom day | 5.47<br>(3.16-7.76) | -2.40<br>(-4.75- -0.06) | 0.056 |
|  | 1<sup>st</sup> day of symptom | 3.07<br>(0.88-5.22) | One day before 1<sup>st</sup> symptom day | 2.92<br>(0.50-5.33) | 0.15<br>(-2.20- 2.37) | 0.91 |
| Acrophase | One day after 1<sup>st</sup> symptom day | -2.30<br>(-2.60- -2.00) | Asymptomatic | -2.45<br>(-2.50- -2.39) | 0.14<br>(-0.15- 0.44) | 0.33 |
|  | One day before 1<sup>st</sup> symptom day | -2.52<br>(-3.31- -1.71) | Asymptomatic | -2.45<br>(-2.50- -2.39) | -0.08<br>(-0.79-0.66) | 0.86 |
|  | 1<sup>st</sup> day of symptom | -2.26<br>(-2.73- -1.79) | Asymptomatic | -2.45<br>(-2.50- -2.39) | 0.19<br>(-0.24- 0.63) | 0.36 |
|  | One day before 1<sup>st</sup> symptom day | -2.52<br>(-3.31- -1.71) | One day after 1<sup>st</sup> symptom day | -2.30<br>(-2.60- -2.00) | -0.22<br>(-1.11-0.70) | 0.63 |
|  | 1<sup>st</sup> day of symptom | -2.26<br>(-2.73- -1.79) | One day after 1<sup>st</sup> symptom day | -2.30<br>(-2.60- -2.00) | 0.04<br>(-0.36-0.46) | 0.86 |
|  | 1<sup>st</sup> day of symptom | -2.26<br>(-2.73- -1.79) | One day before 1<sup>st</sup> symptom day | -2.52<br>(-3.31- -1.71) | 0.26<br>(-0.40-0.92) | 0.41 |

## DISCUSSION

In this prospective study, longitudinally evaluated HRV metrics were found to be associated with a positive SARS-CoV-2 diagnosis and COVID-19 symptoms. Significant changes in these metrics were observed 7 days prior to the diagnosis of COVID-19. To the best of our knowledge this is the first study to demonstrate that physiological metrics derived from a commonly worn wearable device (Apple Watch) can identify and predict SARS-CoV-2 infection prior to diagnosis with a SARS-CoV-2 nasal PCR swab. These preliminary results identify a novel easily measured physiological metric which may aid in the tracking and identification of SARS-CoV-2 infections.

Current means to control COVID-19 spread rely on case isolation and contact tracing, which have played a major role in the successful containment of prior infectious disease outbreaks.^18-20^ However, the variable incubation period, high percentage of asymptomatic carriers, and infectivity during the pre-symptomatic period of COVID-19 have made containment challenging.^21^ This has further limited the utility of systematic screening technologies reliant on vital sign assessment or self-reporting of symptoms.^7^ Advances in digital health provide a unique opportunity to enhance disease containment. Wearable devices are commonly used and well accepted for health monitoring.^9,22^ Commercially available devices are able to continually collect several physiological parameters. Unlike App-based platforms, wearable devices have the advantage of not requiring users to actively participate aside from regular use of the device. Prior to the COVID-19 pandemic, population level data from the Fitbit wearable device demonstrated effectiveness at real-time geographic surveillance of influenza-like illnesses through the assessment of physiological parameters.^23^ This concept was recently expanded during the COVID-19 pandemic by Quer and colleagues who demonstrated that the combination of symptom-based data with resting heart rate and sleep data from wearable devices was superior to relying on symptom-based data alone to identify COVID-19 infections.^10^

HRV has been shown to be altered during illnesses with several small studies demonstrating changes in HRV associated with and predictive of the development of infection.^24^ Ahmad and colleagues followed 21 subjects undergoing bone marrow transplant finding a significant reduction in root mean square successive difference metrics prior to the clinical diagnosis of infection. Furthermore, wavelet HRV was noted to decrease by 25% on average 35 hours prior to a diagnosis of sepsis in 14 patients.^16^ In another study in 100 infants, significant HRV changes were noted 3-4 days preceding sepsis or systemic inflammatory response syndrome with the largest increase being seen 24 hours prior to development.^15^ Building on these observations demonstrating that ANS changes accompany or precede infection, our team launched the Warrior Watch Study.

We demonstrated that significant changes in the circadian pattern of HRV, specifically SDNN’s amplitude, was associated with a positive COVID-19 diagnosis. Interestingly, when we compared these changes over the seven days preceding the diagnosis of COVID-19 we continued to see significant alterations in amplitude when compared to individuals without COVID-19. This demonstrates the predictive ability of this metric to identify infection. Interestingly when we follow individuals 7-14 days after diagnosis with COVID-19, we find that the circadian HRV pattern starts to normalize and is no longer statistically different from an uninfected pattern. As an exploratory analysis we evaluated how HRV was impacted by symptoms associated with a COVID-19 diagnosis, since individuals may not be tested despite symptoms. We found significant changes in the amplitude of the circadian HRV pattern on the first day of symptoms, with a trend toward statistical significance on the day before and after symptoms are reported. Taken together, these findings highlight the possible use of HRV collected via wearable devices to identify and predict COVID-19 infections.

There are several limitations to our study. First, there was a small number of participants who were diagnosed with COVID-19 in our cohort limiting our ability to determine how predictive HRV can be of infection. However, these preliminary findings support the further evaluation of HRV as a metric to identify and predict COVID-19 and warrant further study. An additional limitation is the sporadic collection of HRV by the Apple Watch. While our statistical modelling was able to account for this a denser dataset would allow for expanded evaluation of the relationship between this metric and infections/symptoms. The Apple Watch also only provides HRV in one time-domain (SDNN), limiting assessment of the relationship between other HRV parameters with COVID-19 outcomes. Lastly, an additional limitation is that we relied on self-reported data in this study, precluding independent verification of COVID-19 diagnosis.

In summary, we demonstrated a relationship between longitudinally collected HRV acquired from a commonly used wearable device and SARS-CoV-2 infection. These preliminary results support the further evaluation of HRV as a biomarker of SARS-CoV-2 infection by remote sensing means. While further study is needed, this may allow for the identification of SARS-CoV-2 infection during the pre-symptomatic period, in asymptomatic carriers and prior to diagnosis by a SARS-CoV-2 nasal PCR tests. These findings warrant further evaluation of this approach to track and identify COVID-19 infections and possibly other type of infections.

## Data Availability

Data collected for this study will not be made publicly available.

## Acknowledgement

Support for this study was provided by the Ehrenkranz Lab For Human Resilience, the BioMedical Engineering and Imaging Institute, The Hasso Plattner Institute for Digital Health at Mount Sinai, The Mount Sinai Clinical Intelligence Center and The Dr. Henry D. Janowitz Division of Gastroenterology.

---

**Supplementary Table 1.** Infection related survey questions.

| Survey Question | Survey Responses |
| --- | --- |
| Have you had any of the following symptoms in the past 24 hours? Check all boxes that apply: | Fever or high temperature, chills, tired or weak, body aches, dry cough, sneezing, runny nose, diarrhea, sore throat, headache, shortness of breath, loss of smell or taste, itchy eyes, none, other [insert], None of these |
| On a scale of 1 - 10, how bad do your symptoms feel today, compared to colds or flu that you have had in the past? | 1 [Extremely mild symptoms], 2, 3, 4, 5, 6, 7, 8, 9, 10 [The worst symptoms that you have ever had] |
| If you have been tested for the Coronavirus by nasal PCR since starting this study, what was the result of your test: | Positive / Negative / I have not been tested for Coronavirus/Prefer not to say |
| If you were tested for the Coronavirus by nasal PCR since starting the study, what is the date you were tested? | Date: _____ |
| If you have been tested for the Coronavirus by blood antibody since starting this study, what was the result of your test: | Positive / Negative / I have not been tested for Coronavirus/Prefer not to say |
| If you were tested for the Coronavirus by blood antibody since starting the study, what is the date you were tested? | Date: _____ |

**Supplementary Figure 1.**
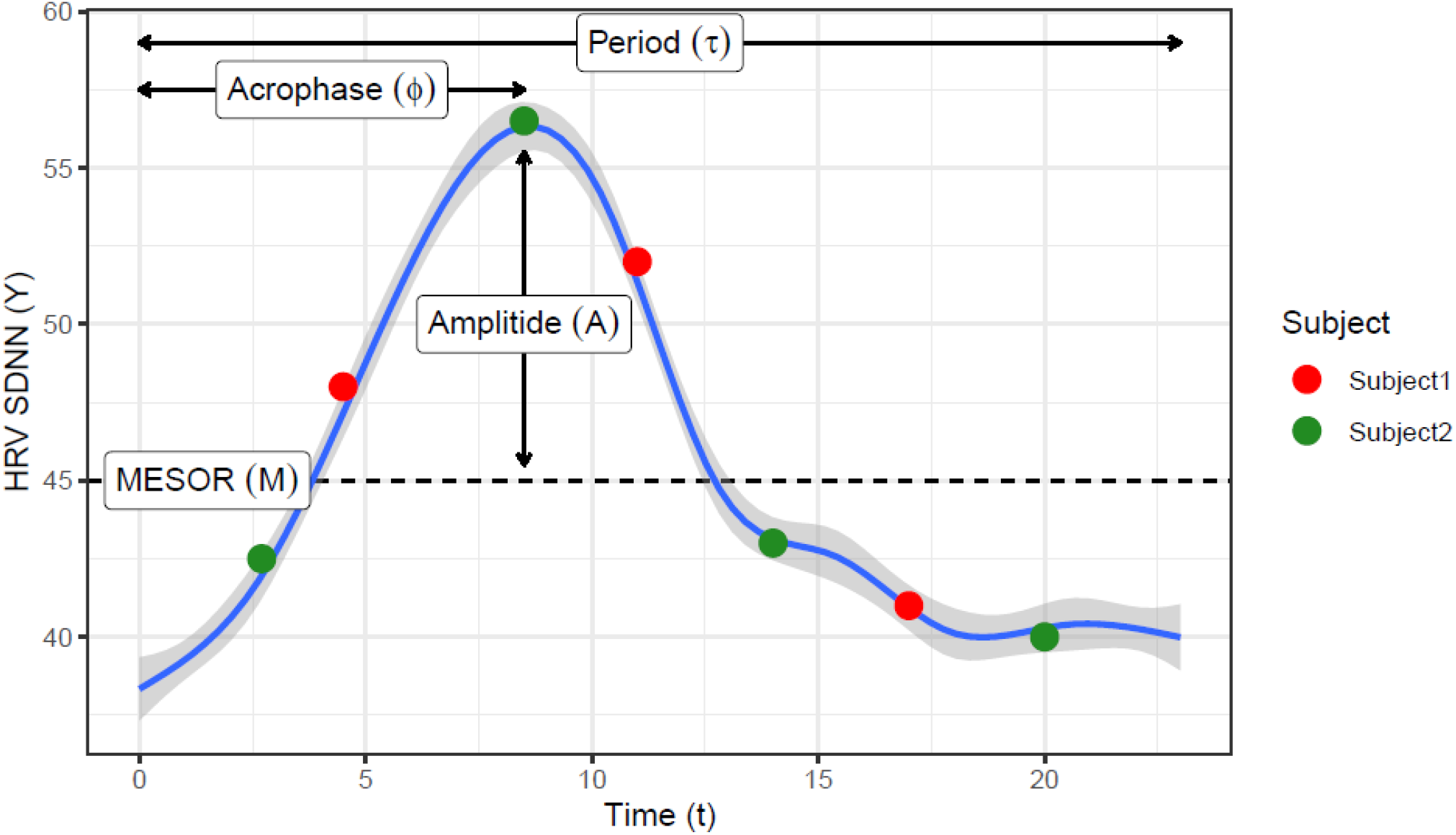
Locally estimated scatterplot smoothing (loess) curve showing a daily circadian pattern on HRV measures. Such pattern can be represented by the COSINOR model using 3 parameters: the rhythm-adjusted mean (MESOR), half the extent of variation within a day (Amplitude) and the time of overall high values recurring in each day (acrophase). Red and green dots represent hypothetical sampling times though the day from two subjects that have the same daily curve, showing that features like maximum, range, or CV will be easily biased by the sampling time.

